# Computer-Aided Reading of Chest Radiographs for Pediatric Tuberculosis: Current Status and Future Directions

**DOI:** 10.1101/2024.10.08.24314837

**Authors:** Mackenzie DuPont, Robert Castro, Sandra V. Kik, Megan Palmer, James A. Seddon, Devan Jaganath

**Affiliations:** Division of Pediatric Emergency Medicine, Baylor College of Medicine, Houston, USA; Division of Pulmonary and Critical Care Medicine, University of California, San Francisco, USA; Center for Tuberculosis, Department of Medicine, University of California, San Francisco, USA; FIND, Geneva, Switzerland; Desmond Tutu TB Centre, Department of Paediatrics and Child Health, Stellenbosch University, Cape Town, South Africa; Department of Infectious Disease, Imperial College London, London, UK; Division of Pediatric Infectious Diseases, University of California, San Francisco, USA

**Author notes:** <u>Corresponding Author</u> Devan Jaganath, MD, University of California, San Francisco, 550 16th St., 4th Floor, San Francisco, CA 94158 USA.

## Abstract

Computer-aided detection (CAD) systems for automated reading of chest X-rays (CXRs) have been developed and approved for tuberculosis (TB) triage in adults, but not in children. However, CXR is frequently the only adjunctive tool to clinical assessment for the evaluation of pediatric TB in primary health settings, and children would benefit from CAD models that can detect their unique clinical and radiographic features. The current literature on CAD CXR algorithms for detecting TB and other pulmonary infections in children is limited, but highlights promising models and approaches. To advance CAD for childhood TB, large, diverse, pediatric CXR datasets are required that are linked to standardized clinical and radiographic TB classifications. These datasets could be used to train or fine-tune existing algorithms for TB screening, diagnosis and severity stratification. It is critical to include children in CAD models to increase equity and reduce the global burden of TB disease.

## INTRODUCTION AND CONTEXT

At the recent United Nations high level meeting on tuberculosis (TB), there was a commitment to prevent and care for 90% of individuals with TB. This goal will be challenging unless innovative tools are developed to improve TB diagnosis for children. In particular, computer aided detection (CAD) systems that utilize artificial intelligence for automated chest x-ray (CXR) reading have been limited to adults and need to be expanded to children.

CXR remains an important part of the recommended evaluation for pediatric TB, but there are challenges to implementation.^1,2^ The radiographic spectrum of TB disease in children varies widely,^3^ and includes hilar lymphadenopathy, consolidation, airway compression, pleural effusion and cavitation. Moreover, accuracy and inter-reader reliability to interpret CXRs has been sub-optimal,^4,5^ and it can be technically challenging to obtain high-quality CXRs in young children.^6^

In general, CAD systems to detect CXR abnormalities follow four steps: 1) image preprocessing and enhancement, 2) extraction of the region of interest such as through lung segmentation, 3) identification of key features and 4) image classification.^7^ The majority of current CAD systems use deep learning algorithms;^8^ this generally requires large data sets for training, though this can be partially mitigated using pre-trained ensemble convolutional neural networks (CNNs) and transfer learning to fine-tune existing models.^8^

In 2021, World Health Organization (WHO) recommended that CAD programs may be used in place of human readers for interpreting CXRs for both TB screening and triage in individuals aged 15 years and older.^1^ There are also several efforts to increase access to portable or mobile CXR machines that can run CAD algorithms with minimal processing requirements.^9,10^ Despite this progress, only a few CAD systems have been developed for children below 15 years.^11^

To guide the advancement of CAD systems for children, we reviewed the current evidence for children evaluated for TB and pneumonia, and discussed the barriers and considerations for future model development.

## SEARCH STRATEGY AND SELECTION CRITERIA

We examined the literature on CAD for tuberculosis and pneumonia in children. We included pneumonia as children with TB can initially present with acute pneumonia symptoms,^12^ and for the potential to adapt these tools for TB. We searched PubMed and Google Scholar using the terms “computer aided detection,” “computer aided diagnosis,” computer aided,” “artificial intelligence,” “machine learning,” and “deep learning,” with “tuberculosis” and/or “pediatric” to identify any published study in English until February 2024 that utilized computer software to analyze CXRs for pulmonary pathology detection in children. Although there is growing work in other imaging modalities, we limited this summary to CXR because it is the current standard for TB evaluation and the focus of CAD algorithms for TB in adults. The identified articles underwent full text review including citations to yield the final reference list based on originality and relevance.

### CURRENT EVIDENCE ON CAD SYSTEMS FOR CHILDHOOD TB AND PNEUMONIA

#### CAD systems for childhood TB

We identified two published studies on the accuracy of CAD for children with signs and symptoms of TB (**Table 1**). Mouton et al. investigated the use of CAD to detect lung abnormalities from the CXRs of 119 children 0-5 years old presenting to the emergency unit with TB symptoms in South Africa.^13,14^ The authors performed lung field segmentation and subdivision, feature extraction, and k-nearest neighbors (kNN) classification to determine if a given lung region was abnormal based on radiologist annotation.^13,15,16^ They found that the area under the receiver operating characteristic curve (AUC) was 0·78 when averaged across 42 lung regions.^13^ However, the classification of abnormalities in the basal and peri-hilar regions of the lungs performed poorly. In the second study, Palmer et al. examined the performance of the commercial CAD algorithm CAD4TB version 7.0 (Delft Systems, ‘s-Hertogenbosch, The Netherlands) among South African children less than 13 years old with signs and symptoms of pulmonary TB.^17^ Based on expert human readers, the AUC of CAD4TB was 0·58 among 80 children to classify a CXR as consistent with TB. When 445 separate CXRs were used to fine-tune the algorithm, the AUC improved to 0·72. Accuracy based on Confirmed vs. Unlikely TB was also evaluated in 53 children, and AUC improved from 0·68 to 0·78 after fine-tuning. Strengths of this study included the use of standard case definitions for childhood TB classification and the inclusion of multiple CXR readers. While the test set was small and accuracy below the WHO target accuracy for a TB triage test (≥90% sensitivity, ≥70% specificity),^18^ this study emphasizes the potential to improve current CAD algorithms for children.

**Table 1.**
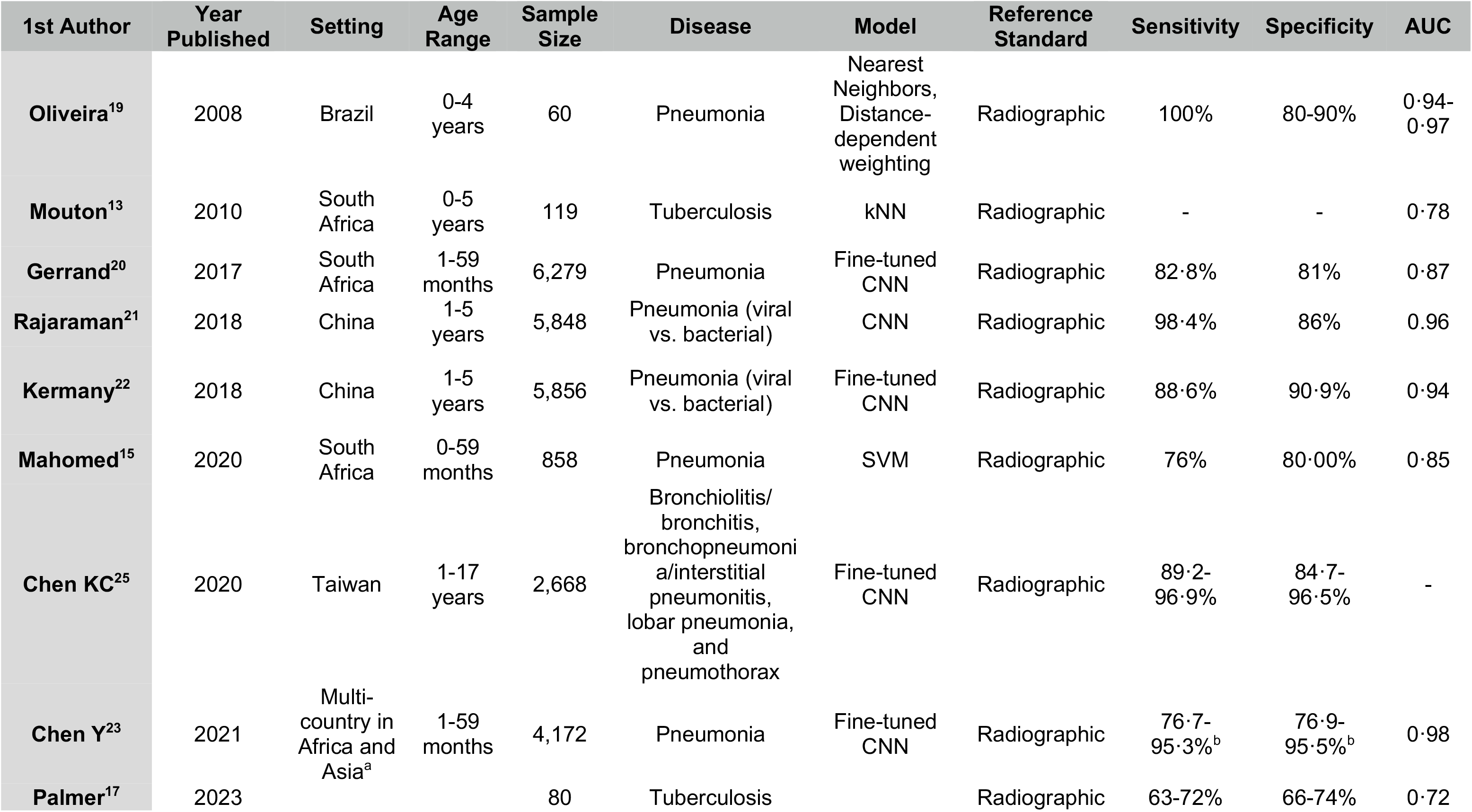
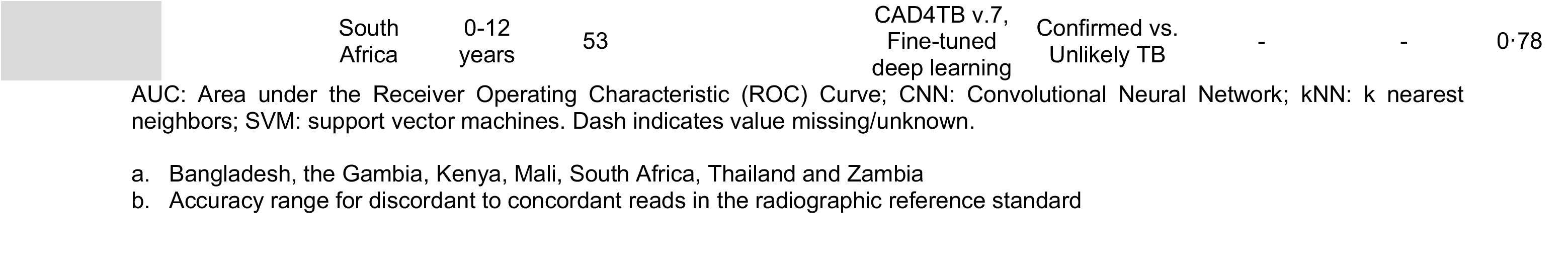
Publications on the use of CAD for pediatric chest x-rays for tuberculosis and pneumonia.

Several commercially available CAD algorithms for the detection of pulmonary TB indicate that they can be used for children.^11^ The CAD product with the lowest age indication is Radify (2 years and older, Envisionit, Johannesburg, South Africa), followed by 3 years and older for qXR (qure.AI, Mumbai, India), 4 years and older for CAD4TB, and 10 years and older for JLD-02K (JLK, Seoul, South Korea).^11^ However, no published estimates of performance in children have been found, neither through this literature review nor by a review of all publications listed on the AI4HLTH.org website or on the products’ respective websites.^11^

#### CAD systems for childhood pneumonia

One of the earliest CAD systems for pediatric pneumonia was called Pneumo-CAD.^19^ The model was developed from a database of 40 CXRs of children (20 with radiographic classification of bacterial pneumonia) from Brazil who presented to a hospital with symptoms of pneumonia. Two wavelet-based approaches were performed to differentiate a set of images with and without pneumonia. Based on a radiographic reference standard, the AUC ranged from 0·94-0·97 on a test set of 20 CXRs, with a sensitivity of 100% and specificity from 80-90%.^19^ The main limitation was a small dataset with 74% pneumonia prevalence, which could lead to overestimation of accuracy.

Several deep learning CAD models have been created for pediatric pneumonia. First, Gerrand et al. fine-tuned a CNN model with transfer learning,^20^ utilizing a pneumococcal conjugate vaccine (PCV)-13 CXR dataset derived from a case-control study in South Africa of hospitalized HIV-uninfected children between one and 59 months old. It contained 3,905 AP CXRs, including 1,394 with pneumonia based on a radiographic reference standard. The algorithm was assessed with five-fold cross-validation, and the authors achieved an AUC of 0·87 with a sensitivity of 83% and a specificity of 81%.^20^ However, the use of healthy controls without lower respiratory tract infections may have overestimated performance. Another study was conducted by Rajaram et al. who tested four types of CNNs to differentiate between normal versus viral versus bacterial pneumonia based on expert read, on a dataset of 5,848 pediatric AP CXRs of children aged one to five years from Guangzhou Women and Children’s Medical Center in China.^16,21^ A customized Visual Geometry Group (VGG)-16 model achieved 98% sensitivity and 86% specificity to classify viral versus bacterial pneumonia.^21^ With the same dataset from China, Kermany et al. fine-tuned a CNN model first developed to classify age-related macular degeneration and diabetic macular edema.^22^ The authors used 5,232 pediatric CXRs to train their model and thereafter used 624 other pediatric CXRs to validate it. Their CAD model was able to reach 89% sensitivity and 91% specificity in the validation dataset to discriminate viral from bacterial pneumonia on CXR.^22^

The Pneumonia Etiology Research for Child Health (PERCH) study was a multi-center initiative from seven countries in Africa and Asia to assess the causes of pneumonia in hospitalized children under 5 years with a clinical presentation of severe or very pneumonia. Two CAD systems were developed using CXRs from this study to detect the WHO-defined radiographic primary-endpoint of pneumonia.^15,23^ First, CAD4Kids was a model developed from a subset of 858 CXRs of children from South Africa.^15^ The authors used lung field segmentation, feature extraction using texture analysis and support vector machines to classify pneumonia by CXR.^15^ Compared to radiologist consensus, CAD4Kids demonstrated an AUC of 0·85 (95% CI 0·82-0·88) with a sensitivity of 76% and specificity of 80% using ten-fold cross validation. A strength of this model was that it was able to identify abnormalities in the peri-hilar region.^13,15^ The authors also assessed accuracy by HIV status, and found that the AUC was overall similar in HIV-infected children (0·81) versus children who were HIV-exposed uninfected (0·85) and HIV-unexposed (0·845). The second CAD system used transfer learning based on a CNN model that was developed from a primarily adult CXR data set from the United States (CheXpert, N = 224,316) followed by fine-tuning on a larger number of PERCH CXRs (N=4,172).^23^ Utilizing 410 images from two different WHO image sets for validation, the model had an AUC of 0·977 (95% CI 0·97-0·98) for pneumonia, and could achieve 95·3% sensitivity and 95·5% specificity. Notably, among CXRs with low agreement between human readers, the accuracy reduced with an AUC of 0·85 (95% CI 0·82-0·87), sensitivity of 76·7% and specificity of 76·9%.^23^ A limitation of these studies was that the WHO pneumonia definitions were designed to evaluate vaccine efficacy and are not used for patient care .^24^ Moreover, as children were hospitalized with severe or very severe pneumonia, their abnormal CXR findings may have been more easily identifiable than in an outpatient setting where children have less severe disease.

KC Chen et al. moved beyond a single disease process to detect pneumonia as well as bronchiolitis/bronchitis, and pneumothorax, in hospitalized children 1-17 years old from Taiwan.^25^ They fine-tuned a CNN with 2,137 CXRs, and evaluated accuracy with 531 CXRs based on a radiographic reference standard. The model achieved 96·9% sensitivity and 96·5% specificity in detecting lobar pneumonia from normal images.^25^ Detection of the other conditions were slightly lower, with sensitivities ranging from 89%-92%, and specificities ranging from 85%-96%. The comparison was to normal images which again may have overestimated performance. They also noted that the levels of disagreement between the model and the radiographic reference were highest for children under five years, and particularly those with peri-hilar changes.

### LESSONS LEARNED AND ONGOING NEEDS

We found few studies on CAD systems for the detection of pediatric TB and pneumonia. The current evidence highlights that deep learning models achieved moderate to good accuracy to classify childhood pneumonia, and fine-tuning through transfer learning has partially mitigated the need for massive image datasets for training. At the same time, several of the described pediatric CAD models had difficulty with images from young children and with abnormalities in the peri-hilar region. The few publications specifically on CAD for childhood TB have shown lower accuracy than that of pneumonia models, but have been studied in smaller datasets and further fine-tuning was shown to improve performance. Overall, these studies can inform key next steps to support the development of accurate algorithms for childhood TB.

First, pediatric CXR repositories are needed with several unique characteristics (**Box 1**). The CXRs need to reflect the range of specific features seen in childhood TB, as well as the normal physical and anatomical developmental changes in children across the age spectrum, such as the presence or absence of the thymus. Given that adolescents have adult-type disease that likely could be evaluated by current CAD systems, these repositories should prioritize CXRs from children less than 10 years old. CXRs should represent a diverse geographic distribution to reflect differences in co-morbidities, other non-TB respiratory conditions, and variation in image acquisition techniques and quality to reflect real-world CXRs. The associated clinical data should support key sub-group analyses such as for infants, children living with HIV and those with malnutrition. CXRs that are used for training of the CAD system should ideally be interpreted by experts who can distinguish key features of pulmonary TB using standardized definitions.^26,27^ Lateral-view CXRs can be helpful for evaluation of hilar lymphadenopathy in children, but current commercial CAD algorithms have not been developed to interpret this view and should be prioritized. Different image repositories may be needed depending on the intended use-case of the CAD product, whether for TB screening of asymptomatic children, for differentiating pulmonary TB from other respiratory diseases in a symptomatic child, or to distinguish children with severe or non-severe TB to inform the appropriate treatment regimen and duration. These image sets should be made publicly available to allow new algorithms to be transparently developed.

In addition, the lack of a clear gold standard diagnostic test for pediatric TB is a barrier for CAD development and validation. Children have paucibacillary disease, and using a microbiological reference standard will exclude a large proportion of children who have unconfirmed, but clinically-diagnosed TB, and bias the population to a more severe disease phenotype. At the same time, a clinical reference standard may incorporate CXR findings in the diagnosis of TB and also bias performance estimates. Using both reference standards provides an accuracy range while recognizing their limitations. The performance of any CAD algorithm should also be compared to a radiographic reference standard read by expert clinicians and/or radiologists to assess their accuracy and reliability as a replacement to a human-interpreted CXR. For all analyses, accuracy reporting should be standardized to support comparison between CAD algorithms. This includes presenting findings aligned with the WHO target product profile for a TB triage test,^18^ such as the specificity at 90% sensitivity.

The desired output of a CAD algorithm for children also needs to be further defined. Given the heterogenous CXR findings, it may not be possible to obtain an algorithm that can detect all forms of childhood pulmonary TB. Instead, an output of likely abnormalities may have greater value to guide confirmatory testing for TB or other lower respiratory diseases. Even classifying abnormal from normal CXRs could be important in primary care settings to guide next steps in evaluation. A CAD algorithm that could distinguish severe versus non-severe TB disease may also guide which children are eligible for shorter TB regimens.

Lastly, it is important to consider implementation and infrastructure needs. Greater access to digital x-rays and technical training can improve image quality and potentially the performance of the CAD algorithms. Current ultra-portable x-ray machines that support CAD algorithms are promising,^28^ but have not yet been assessed in children. In addition, models should be integrated into existing treatment decision algorithms for childhood TB to guide new approaches in clinical decision-making. Further work is needed to assess how the resources to implement CAD will improve TB detection in children as compared to other triage and diagnostic approaches. This will be influenced by when and where CAD is utilized along the diagnostic pathway, such as during contact screening or in a health facility. While a separate pediatric CAD algorithm may be needed, validated models should be incorporated into existing adult CAD products to encourage equitable access and scale up of TB detection in individuals of all ages.

## CONCLUSIONS

There have been advances in the use of automated CXR reading to detect pulmonary disease in children. Collaboration between developers and childhood TB researchers is critical to pool data and expertise to support the development and evaluation of novel pediatric TB CAD systems. Ultimately this would enable health providers in low-resource settings to make faster, informed clinical decisions to reduce delays in diagnosis and care for children with TB.

## Data Availability

Relevant data from the referred articles are indicated in the manuscript, and cited.

## CONTRIBUTIONS

The work was conceptualized by MD, SVK, MP, JAS, and DJ. The data review was completed by MD, RC, and DJ. Writing was completed by MD, RC, and DJ, with review and editing by all authors.

## ACKNOWLEDGEMENT

There was no funding source for this work.

---

**BOX 1.**
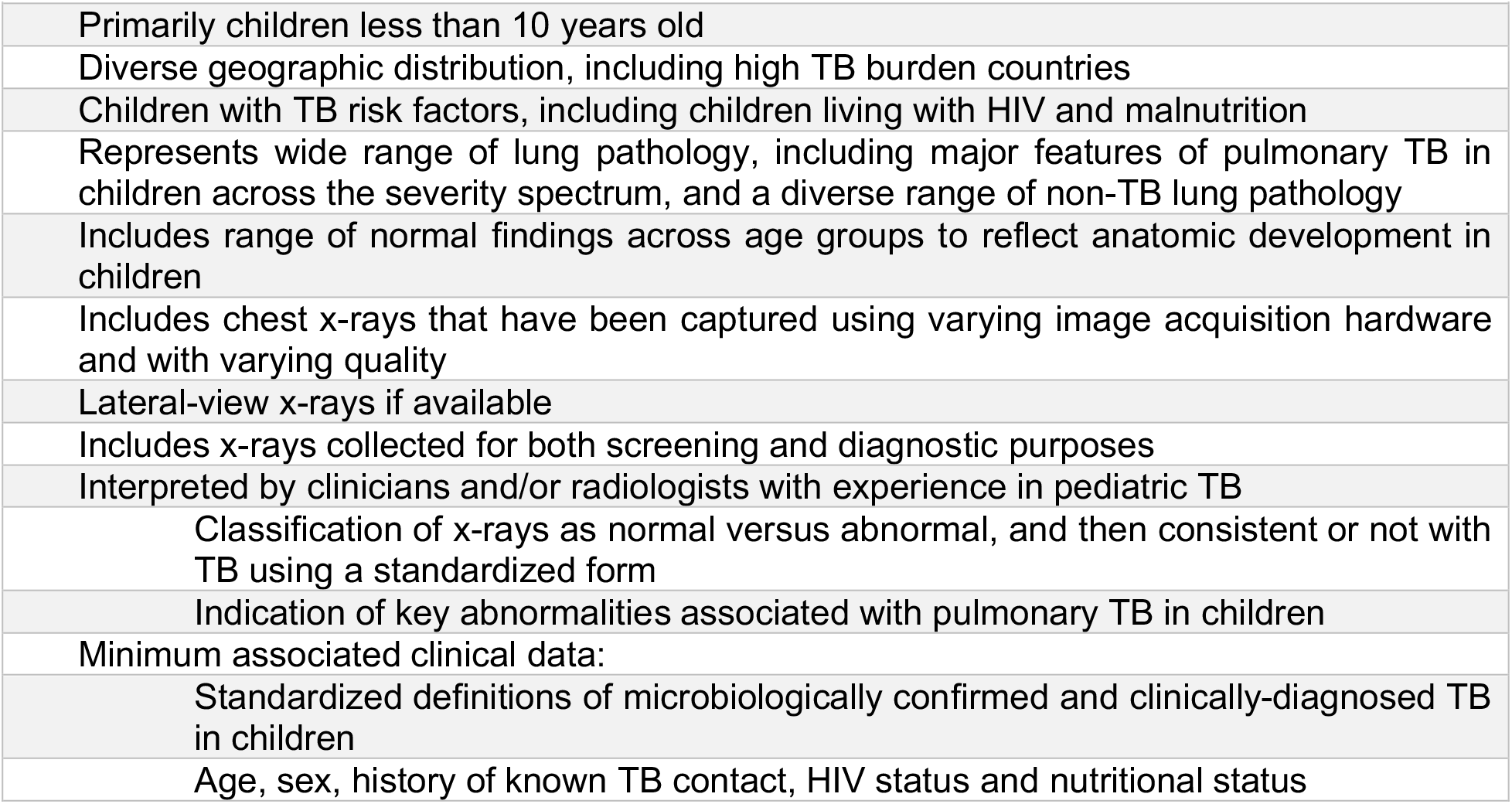
Ideal characteristics for a chest X-ray repository for childhood tuberculosis to
support CAD product development.

## REFERENCES

1. World Health Organization. WHO consolidated guidelines on tuberculosis. Module 2: Screening - Systematic screening for tuberculosis disease. Geneva, World Health Organization. 2021.

2. World Health Organization. Guidance for national tuberculosis programmes on the management of tuberculosis in children. WHO: Geneva, 2014.

3. Marais BJ, Gie RP, Schaaf HS, et al. A proposed radiological classification of childhood intra-thoracic tuberculosis. Pediatr Radiol 2004; 34(11): 886–94.

4. Seddon JA, Padayachee T, Du Plessis AM, et al. Teaching chest X-ray reading for child tuberculosis suspects. Int J Tuberc Lung Dis 2014; 18(7): 763–9.

5. Brinkmann F, Hogrefe J, Berthold L-D, Müller-Heine A, Hansen G, Schwerk N. Reliability of radiological diagnosis in children and adolescents with confirmed tuberculosis infection. European Respiratory Journal 2013; 42(Suppl 57): P4481.

6. Thukral BB. Problems and preferences in pediatric imaging. Indian J Radiol Imaging 2015; 25(4): 359–64.

7. Ueda D, Shimazaki A, Miki Y. Technical and clinical overview of deep learning in radiology. Japanese Journal of Radiology 2019; 37(1): 15–33.

8. Beam AL, Kohane IS. Big Data and Machine Learning in Health Care. Jama 2018; 319(13): 1317–8.

9. FIND. Digital Chest Radiography and Computer-Aided Detection (CAD) Solutions for Tuberculosis Diagnostics: Technology Landscape Analysis. FIND: Geneva, 2021.

10. Melendez J, Philipsen R, Chanda-Kapata P, Sunkutu V, Kapata N, van Ginneken B. Automatic versus human reading of chest X-rays in the Zambia National Tuberculosis Prevalence Survey. Int J Tuberc Lung Dis 2017; 21(8): 880–6.

11. STOP TB and FIND. AI4Hlth. 2021. https://www.ai4hlth.org/.

12. Oliwa JN, Karumbi JM, Marais BJ, Madhi SA, Graham SM. Tuberculosis as a cause or comorbidity of childhood pneumonia in tuberculosis-endemic areas: a systematic review. Lancet Respir Med 2015; 3(3): 235–43.

13. Mouton A, Pitcher RD, Douglas TS. Computer-aided detection of pulmonary pathology in pediatric chest radiographs. Med Image Comput Comput Assist Interv 2010; 13(Pt 3): 619–25.

14. Tezoo T, Douglas TS. Interactive segmentation of airways from chest x-ray images using active shape models. 2012 Annual International Conference of the IEEE Engineering in Medicine and Biology Society; 2012: IEEE; 2012. p. 1498–501.

15. Mahomed N, van Ginneken B, Philipsen R, et al. Computer-aided diagnosis for World Health Organization-defined chest radiograph primary-endpoint pneumonia in children. Pediatr Radiol 2020; 50(4): 482–91.

16. He K, Zhang X, Ren S, Sun J. Deep residual learning for image recognition. Proceedings of the IEEE conference on computer vision and pattern recognition; 2016; 2016. p. 770–8.

17. Palmer M, Seddon JA, van der Zalm MM, et al. Optimising computer aided detection to identify intra-thoracic tuberculosis on chest x-ray in South African children. PLOS Global Public Health 2023; 3(5): e0001799.

18. World Health Organization. High-priority target product profiles for new tuberculosis diagnostics: report of a consensus meeting.2014. https://apps.who.int/iris/bitstream/handle/10665/135617/WHO_HTM_TB_2014.18_eng.pdf?sequence=1 (accessed 9 May 2019.).

19. Oliveira LL, Silva SA, Ribeiro LH, de Oliveira RM, Coelho CJA.SA. Computer-aided diagnosis in chest radiography for detection of childhood pneumonia. Int J Med Inform 2008; 77(8): 555–64.

20. Gerrand J, Williams Q, Lunga D, Pantanowitz A, Madhi S, Mahomed N. Paediatric frontal chest radiograph screening with fine-tuned convolutional neural networks. Annual Conference on Medical Image Understanding and Analysis; 2017: Springer; 2017. p. 850–61.

21. Rajaraman S, Candemir S, Kim I, Thoma G, Antani S. Visualization and Interpretation of Convolutional Neural Network Predictions in Detecting Pneumonia in Pediatric Chest Radiographs. Appl Sci (Basel) 2018; 8(10).

22. Kermany DS, Goldbaum M, Cai W, et al. Identifying Medical Diagnoses and Treatable Diseases by Image-Based Deep Learning. Cell 2018; 172(5): 1122-31.e9.

23. Chen Y, Roberts CS, Ou W, et al. Deep learning for classification of pediatric chest radiographs by WHO’s standardized methodology. PLoS One 2021; 16(6): e0253239.

24. Mahomed N, Fancourt N, de Campo J, et al. Preliminary report from the World Health Organisation Chest Radiography in Epidemiological Studies project. Pediatr Radiol 2017; 47(11): 1399–404.

25. Chen KC, Yu HR, Chen WS, et al. Diagnosis of common pulmonary diseases in children by X-ray images and deep learning. Sci Rep 2020; 10(1): 17374.

26. Palmer M, Gunasekera KS, van der Zalm MM, et al. The diagnostic accuracy of chest radiographic features for pediatric intrathoracic tuberculosis. Clin Infect Dis 2022.

27. Graham SM, Ahmed T, Amanullah F, et al. Evaluation of tuberculosis diagnostics in children: 1. Proposed clinical case definitions for classification of intrathoracic tuberculosis disease. Consensus from an expert panel. J Infect Dis 2012; 205 Suppl 2(Suppl 2): S199–208.

28. Vo LNQ, Codlin A, Ngo TD, et al. Early Evaluation of an Ultra-Portable X-ray System for Tuberculosis Active Case Finding. Trop Med Infect Dis 2021; 6(3).

